# Faecal incontinence is associated with an impaired rectosigmoid brake and improved by sacral neuromodulation

**DOI:** 10.1101/2021.11.30.21266844

**Authors:** Anthony Y. Lin, Chris Varghese, Niranchan Paskaranandavadivel, Sean Seo, Peng Du, Phil Dinning, Ian P. Bissett, Greg O’Grady

## Abstract

**Background and aims:** The rectosigmoid brake, characterized by retrograde cyclic motor patterns on high-resolution colonic manometry has been postulated as a contributor to the maintenance of bowel continence. Sacral neuromodulation (SNM) is an effective therapy for faecal incontinence, but its mechanism of action is unclear. This study aims to investigate the colonic motility patterns in the distal colon of patients with faecal incontinence, and how these are modulated by SNM.

**Methods:** A high-resolution fibre-optic colonic manometry catheter, containing 36 sensors spaced at 1-cm intervals, was positioned in patients with faecal incontinence undergoing Stage 1 SNM. One hour of pre-meal and post-meal recordings were obtained followed by pre- and post-meal recordings with suprasensory SNM. A 700-kcal meal was given. Data were analysed to identify propagating contractions.

**Results:** Fifteen patients with faecal incontinence were analysed. Patients had an abnormal meal response (fewer retrograde propagating contractions compared to controls; p=0.027) and failed to show a postmeal increase in propagating contractions (mean 17 ± 6/h pre-meal vs 22 ± 9/h post-meal, p = 0.438). Compared to baseline, SNM significantly increased the number of retrograde propagating contractions in the distal colon (8 ± 3/h pre-meal vs 14 ± 3/h pre-meal with SNM, p = 0.028). Consuming a meal did not further increase the number of propagating contractions beyond the baseline upregulating effect of SNM.

**Conclusion:** The rectosigmoid brake was suppressed in this cohort of patients with faecal incontinence. SNM may exert a therapeutic effect by modulating this rectosigmoid brake.

**What You Need to Know:** *Background and context:* The rectosigmoid brake, characterized by retrograde propagating motility patterns, has been postulated to contribute to the maintenance of continence. The mechanisms of action of sacral neuromodulation remain inadequately understood and may include modulation of the rectosigmoid brake.

*New findings:* We found that patients with faecal incontinence had an impaired rectosigmoid brake, characterized by a reduced frequency of colonic motor patterns in response to a meal. Rectosigmoid brake activity was upregulated by sacral neuromodulation.

*Limitations:* This was a small cohort of patients with heterogenous faecal incontinence subtypes.

*Impact:* Attenuation of the rectosigmoid brake is a biomarker of faecal incontinence. Rectosigmoid brake responses offer a therapeutic target to evaluate and refine sacral neuromodulation protocols.

**Lay summary:** Patients with faecal incontinence had an attenuated rectosigmoid brake, characterised by fewer postprandial retrograde propagating contractions in the distal colon, however, the rectosigmoid brake function was improved by sacral neuromodulation.

## Introduction

Faecal incontinence (FI) affects between 5-15% of people globally ^1^. It is associated with significant social embarrassment, psychological distress and economic burden ^2,3^. While causes of faecal incontinence are often multifactorial ^4–6^, research has predominantly focused on anorectal physiology with less emphasis on the role of colonic motility.

Sacral neuromodulation (SNM) is an effective treatment for faecal incontinence refractory to medical management ^7^, with sustained long-term benefits ^8^. SNM entails surgical placement of stimulating electrodes adjacent to the sacral nerve root, typically S3 ^9^. Despite its clinical success, the mechanism of action has not been clearly elucidated ^10,11^. A limited understanding of the mechanism of action of SNM has meant no biomarkers exist for monitoring the treatment response to SNM which hinders improvement of the therapy and appropriate patient selection.

SNM has been hypothesised to modulate afferent, central, autonomic, and somatic neural pathways ^11,12^. Chronic SNM may act through somatic afferents to reduce inhibition of sphincter function via ascending central pathways ^11,13^. Locally, external sphincter hypertrophy secondary to stimulation have also been implicated ^11^, however, there is little evidence that SNM affects sphincter activation, anal squeeze pressures, anal reflexes, or internal sphincter slow wave amplitudes ^11,14,15^. Moreover, many patients benefit from SNM despite large sphincter defects ^16,17^, suggesting factors other than the sphincter complex are implicated. Proximal factors such as colonic motility, which is modulated by the sacral nerves, may therefore, also contribute to the pathophysiology of faecal incontinence ^12,18,19^. By stimulating the parasympathetic innervation of the distal colon, SNM may modulate the rectosigmoid brake, a predominantly retrograde, cyclic motor pattern thought to limit rectal filling and contribute to the maintenance of continence ^12,20^, a concept first postulated and demonstrated by Patton et al. ^19^.

In this study, we evaluated the motility of the distal colon in patients with faecal incontinence, and defined how it altered with SNM, using high-resolution colonic manometry (HRCM).

## Methods

Ethical approval was obtained from the New Zealand Health and Disability Ethics Committee (ref: 15/NTA/175). All participants provided informed written consent.

### Eligibility criteria for SNM

Adult patients aged >18 years old who were referred for SNM for FI. Patients were eligible for SNM if they had failed medical management for faecal incontinence and had experienced at least two episodes of faecal incontinence per week for a minimum of 12 months. This was confirmed using a daily bowel diary. All patients underwent a thorough assessment through the pelvic floor clinic at Auckland City Hospital. Baseline incontinence data was assessed via Faecal Incontinence Severity Index (FISI) and a modified Faecal Incontinence Quality of Life scores used to assess clinical response to SNM at Auckland City Hospital ^23^. The decision to perform SNM on each patient was determined by a multidisciplinary pelvic floor team. All included patients provided informed consent.

Patients that were pregnant, suffering severe metabolic, neurologic, or endocrine disorder known to cause colonic dysmotility, previous bowel resection, and/or major lumbosacral injury or malformations were excluded.

### Healthy controls

Control data were amalgamated from a historical cohort ^24^, and an additional two healthy control participants were also recruited. These data were included to compare the meal response between patients with faecal incontinence and healthy controls. These participants received oral mechanical bowel preparation and underwent 2 h of preprandial and postprandial HR colonic manometry recordings, with a 700 kCal meal. A fiber-optic manometry catheter with 72 sensors at 1 cm intervals was used in these patients, though for consistency in comparisons with the preoperative cohort during analysis, only motor events from the most distal 36 sensors (i.e. those located in the descending colon, sigmoid colon, and rectum) were evaluated.

### Interventions

The recordings were taken at the time of the first stage of SNM lead insertion. Colonic manometry recordings were taken during the in-patient stay for the first-stage procedure. More extensive details about the sacral nerve stimulator implant procedure can be found in the **Supplementary Appendix; Methods**. All patients with faecal incontinence were fasted from midnight on the day of the procedure. All patients received a 1 g oral paracetamol and a standardised institutional perioperative analgesia protocol. The choice of performing SNM under sedation or general anaesthesia was left to surgeon preference. Where, under sedation midazolam, remifentanil infusion and/or propofol infusion was used with medications titrated to effect; where general anaesthesia was used, fentanyl and midazolam were given as premedication. The choice of induction agent and neuromuscular blockade was left to the discretion of the anaesthetist. The neuromuscular blockade is typically a short-acting agent, such as rocuronium (clinical duration of 33 min ^25^) to allow for observation of motor responses from test nerve stimulation. Postoperative pain management was standardised and consisted of paracetamol and tramadol or Sevredol. Patients could refuse post-procedural analgesia if they were not needed.

#### Sacral neuromodulation

SNM is typically performed in two-stages. The first stage is a temporary evaluation phase, where in this study cohort all patients had a definitive quadripolar tined lead connected to a temporary stimulator placed surgically. If clinical success was achieved (typically >50% improvement in symptoms per a bowel symptom diary at the end of a 1-month temporary SNM period), patients would then move onto the second stage, wherein a permanent stimulator is placed.

### High-resolution colonic manometry

A fibre-optic HR manometry catheter with 36 sensors at 1-cm intervals was used to measure distal colonic motor activity ^26^. All faecal incontinence patients received one or two Fleet enemas (Fleet Laboratories USA) before surgery to allow for the manometry catheter. Manometry catheter placement was performed at the end of the SNM first-stage procedure. A nylon loop was tied to the tip of the manometry catheter. A Resolution clip (Boston Scientific, Marlborough, MA, USA) was inserted into the working channel of a flexible endoscope to grasp the nylon loop and guide the placement of the manometry catheter per anus. The HR manometry catheter was advanced to a point where the last sensor was no longer visible at the anal verge. Once in position, one or two Resolution clips were used to secure the catheter via the nylon loop to the colon mucosa. A piece of tape was also used to secure the catheter to the buttock. During the recordings, the catheter was connected to a spectral interrogator acquisition unit (FBG-scan 804; FOS&S, Geel, Belgium). A purpose-written LabVIEW program (National Instruments, Austin, Texas, USA) was used to record data.

### Manometry study protocol

The manometry recording commenced once patients were fully awake. Abdominal X-rays were taken approximately 4 h after the surgery to confirm the position of the manometry catheter. First, 2 h of basal recording with no stimulation were performed. After the basal recording, the implanted sacral nerve stimulator lead was connected to a temporary external stimulator, and a further 2 h of recording was undertaken using the standard therapeutic setting (suprasensory level of stimulation), as determined by the Medtronic specialist. The setting was based on a default setting supplied by Medtronic (14 Hz and pulse width of 210 μs). The amplitude was slowly increased in 0.1 V increments until the patient perceived sensory stimulation in the perineum. The final amplitude was set at a level where the patient was aware of the stimulation but remained comfortable. After 2 h of suprasensory stimulation, patients were given a standardised 700 kcal meal, consisting of a chicken sandwich and a Nepro HP drink (Abbott Nutrition, Columbus, OH, USA). While still receiving SNM, patients underwent a further 2 h of recording, after which the HR manometry catheter was disconnected, and the temporary external stimulator was turned off.

Hospital service requirements allowed a subgroup to undergo an extended protocol which involved providing the same 700 kcal meal after surgery, but prior to activating SNM so the baseline meal response pattern of colonic motility could be measured ^23^. Only a subset of patients received this extended protocol (n = 6). Data acquisition methods for the controls have previously been published ^12,20,23^.

### Manometric analysis

Manometric data analysis was performed using a custom-designed software package (PlotHRM; Flinders University). One hour of data from either side of the meal and/or stimulation was extracted for analysis. This was due to unforeseen circumstances such as patients mobilizing and catheter migration that meant recordings beyond 1 hour were heterogeneous. Data for the healthy controls were truncated to equal length to allow for a direct comparison. Event detection and pattern recognition were based on previously described methods and definitions. Propagating contractions were defined as spatiotemporal motor patterns with pressure peaks that occur in four or more adjacent channels (i.e., ≥ 3 cm) and had a trough-to-peak amplitude of ≥ 5 mmHg. All propagating contractions were analyzed with an additional subgroup analysis of the cyclic motor pattern (CMP). Cyclic motor pattern is defined as repetitive propagating contractions with a frequency between 2 and 8 cpm for a duration of 3 min. This is the predominant motor pattern thought to underlie the rectosigmoid brake ^27,28^. Event counts were averaged across multiple subjects by interpolating the data between between sigmoid flexure and rectosigmoid junction of each subject to the centre line of a three-dimensional colonic model generated using data from the Visible Human Project (US National Library of Medicine, Bethesda, Maryland, USA), then projected to the surface of the model as color-maps, as previously described ^29^.

### Statistical analysis

Data are presented as mean (standard error). Nonparametric paired Wilcoxon signed-rank tests were used to compare pre- and postprandial, and pre- and post-stimulation propagating contractions as appropriate. All statistical testing was conducted in R (R Foundation for Statistical Computing, Austria 2014) with p < 0.05 considered statistically significant.

## Results

Overall, 15 patients with faecal incontinence undergoing stage 1 SNM (median age 61, range 45-81; 13 female), of which 6 had postprandial recordings without SNM. Eleven healthy control participants were recruited (median age 52, range 30-69 years; 5 female)

The patient group consisted of 5 patients with urge incontinence, 5 patients with passive incontinence, and 5 patients with mixed incontinence based on pelvic floor assessments. The mean FISI score was 41.3 (range 25-61), and the mean FIQoL score was 70.9 (range 20-95). Further clinical details of patients with faecal incontinence undergoing SNM, are outlined in **Table 1** and **Supplementary Appendix; Table S1**.

**Table 1:** Anorectal physiology results

| Examination | Result |
| --- | --- |
| Endoanal ultrasound* |  |
| Internal sphincter |  |
| Intact | 5 |
| Defect | 6 |
| External sphincter |  |
| Intact | 10 |
| Defect | 1 |
| Anorectal manometry, mmHg** |  |
| Resting | 34.2 ± 19.8 |
| Squeeze | 60.6 ± 35.2 |
| Cough | 55.9 ± 36.8 |

\*Endoanal ultrasound results were not available for three patients. \*\*Anal manometry results were not available for two patients.

Four patients were taking loperamide prior to the operation. No patients received antidiarrheal agents, including loperamide or codeine, on the day of surgery for the duration of hospitalization. On the day of surgery, 11 patients did not receive narcotics postoperatively and 4 patients required more than three doses of either tramadol or Sevredol.

Ten patients underwent SNM placement under sedation while 5 received general anaesthesia. SNM was placed in S3 on all patients, 12 on the left side and 3 on the right side.

### Differences in colonic motility between patients with faecal incontinence and controls

Patients with faecal incontinence had an impaired meal response with respect to the number of total (mean 17 ± 6/h pre-meal vs 22 ± 9/h post-meal, p = 0.438), antegrade (10 ± 3/h pre-meal vs 5 ± 2/h post-meal, p = 0.916) and retrograde (8 ± 3/h pre-meal vs 17 ± 9/h post-meal, p = 0.281) propagating contractions (**Figure 1, 2, 4**). As previously documented, healthy controls showed significant increases in the number of total (21 ± 14/h pre-meal vs 70 ± 12/h post-meal, p < 0.001), antegrade (8 ± 6/h pre-meal vs 18 ± 6/h post-meal, p = 0.014), and retrograde (13 ± 7/h pre-meal vs 52 ± 8/h post-meal, p < 0.001) propagating contractions (**Figure 1, 2 and 4**). Particularly, the magnitude of the delta in retrograde propagating contractions was significantly greater in controls compared to faecal incontinence patients (delta in mean retrograde contractions: 39 vs 9, p = 0.027) (**Figure 2 and 4**).

**Figure 1:**
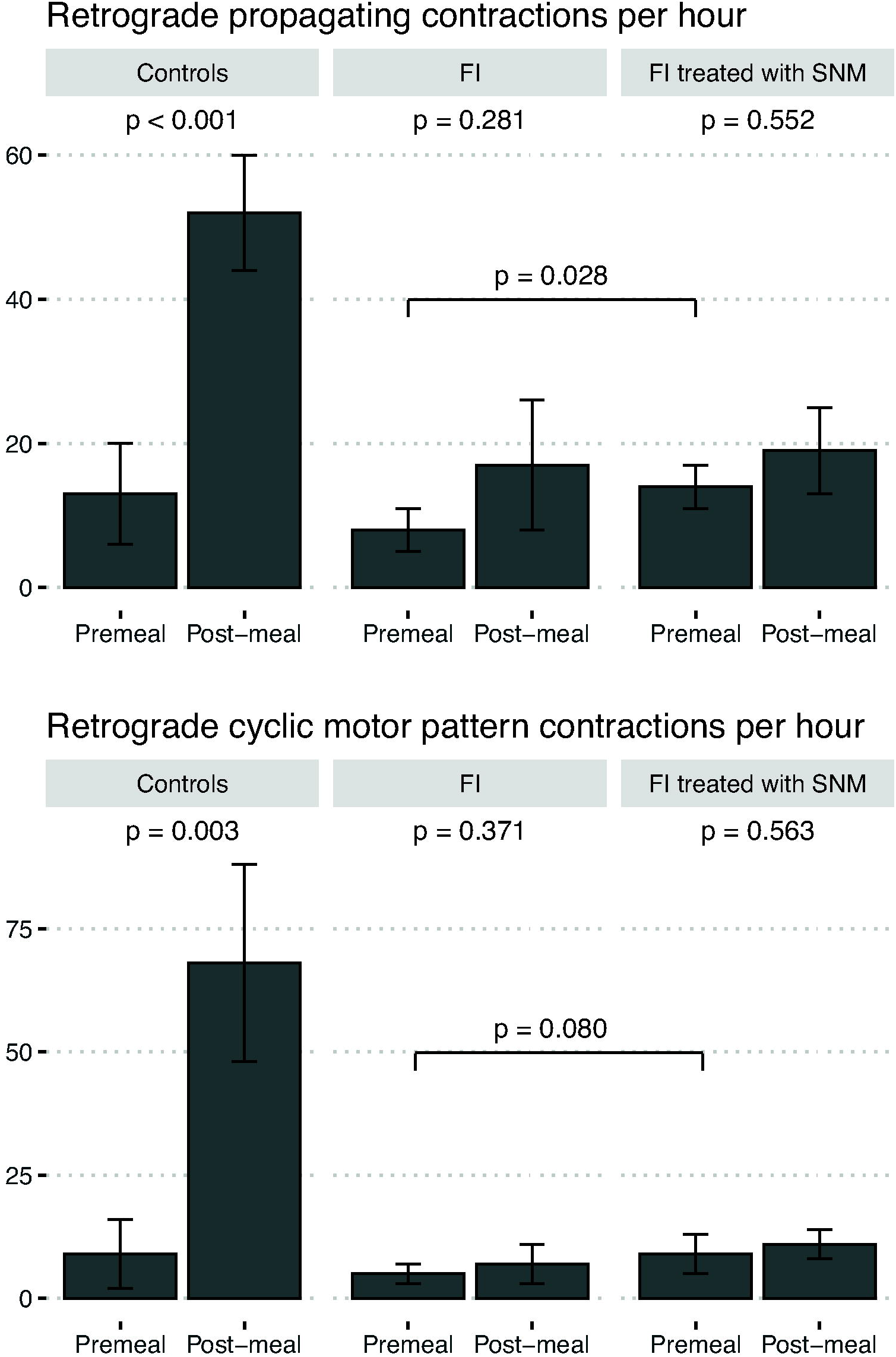
All (top) and cyclic motor pattern-associated (bottom) retrograde propagating contractions in healthy controls, patients with faecal incontinence both before and after sacral neuromodulation. *Plotting mean ± SE

**Figure 2:**
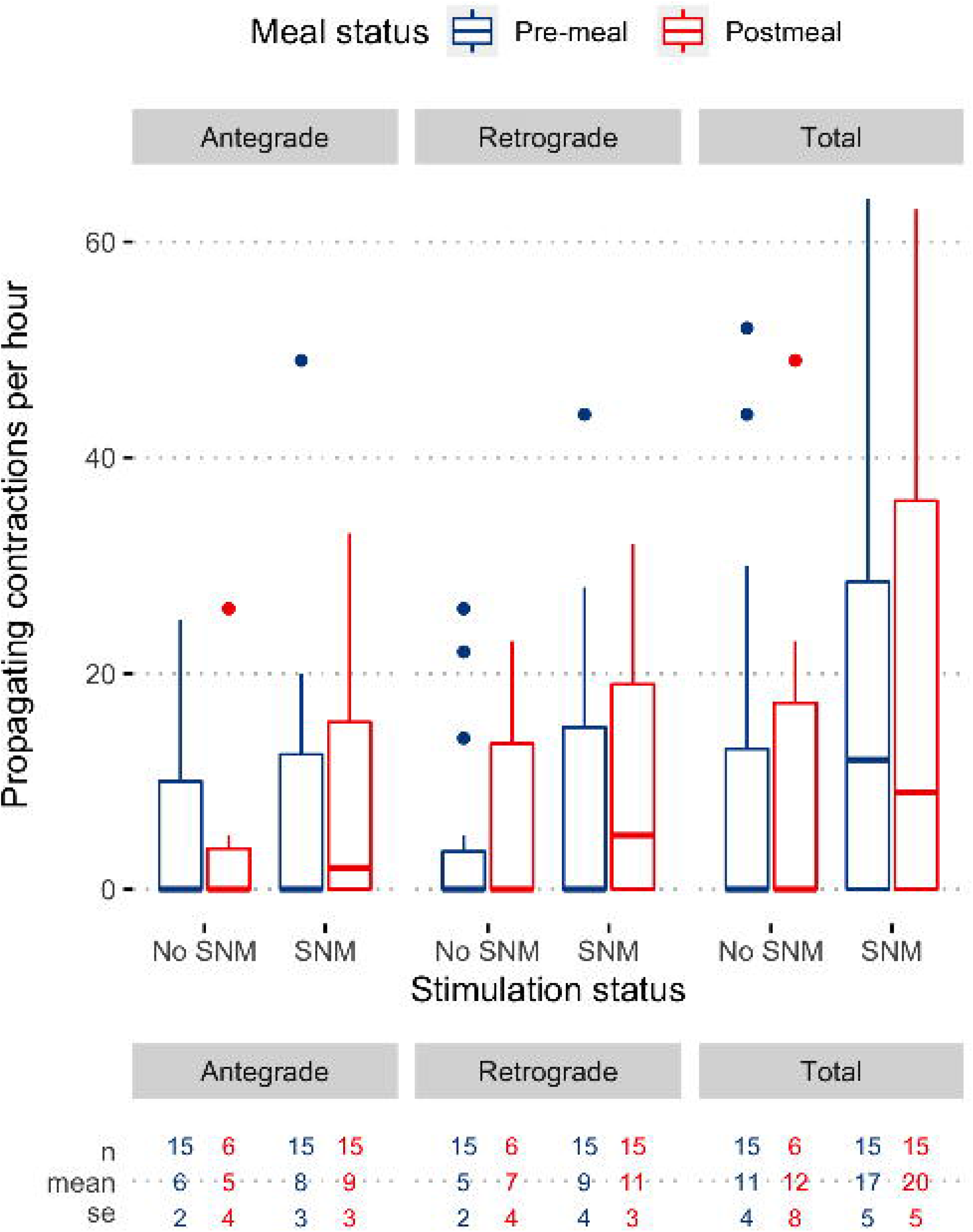
representative examples of pre-meal and postmeal high-resolution colonic manometry data in 10-minute epochs. A: Significant increase in postprandial propagating contraction frequencies in healthy controls; B: Increased postprandial colonic activity but less propagation and decreased activity in faecal incontinence patients compared to controls; C: increase in propagating contractions at baseline and postprandially with SNM in patients with faecal incontinence. SNM appears to increase frequency of propagating events but not to the level seen in the healthy control meal response. *Despite variation in catheter used, only data distal to the splenic flexure were analysed in all cohorts.

**Figure 3:**
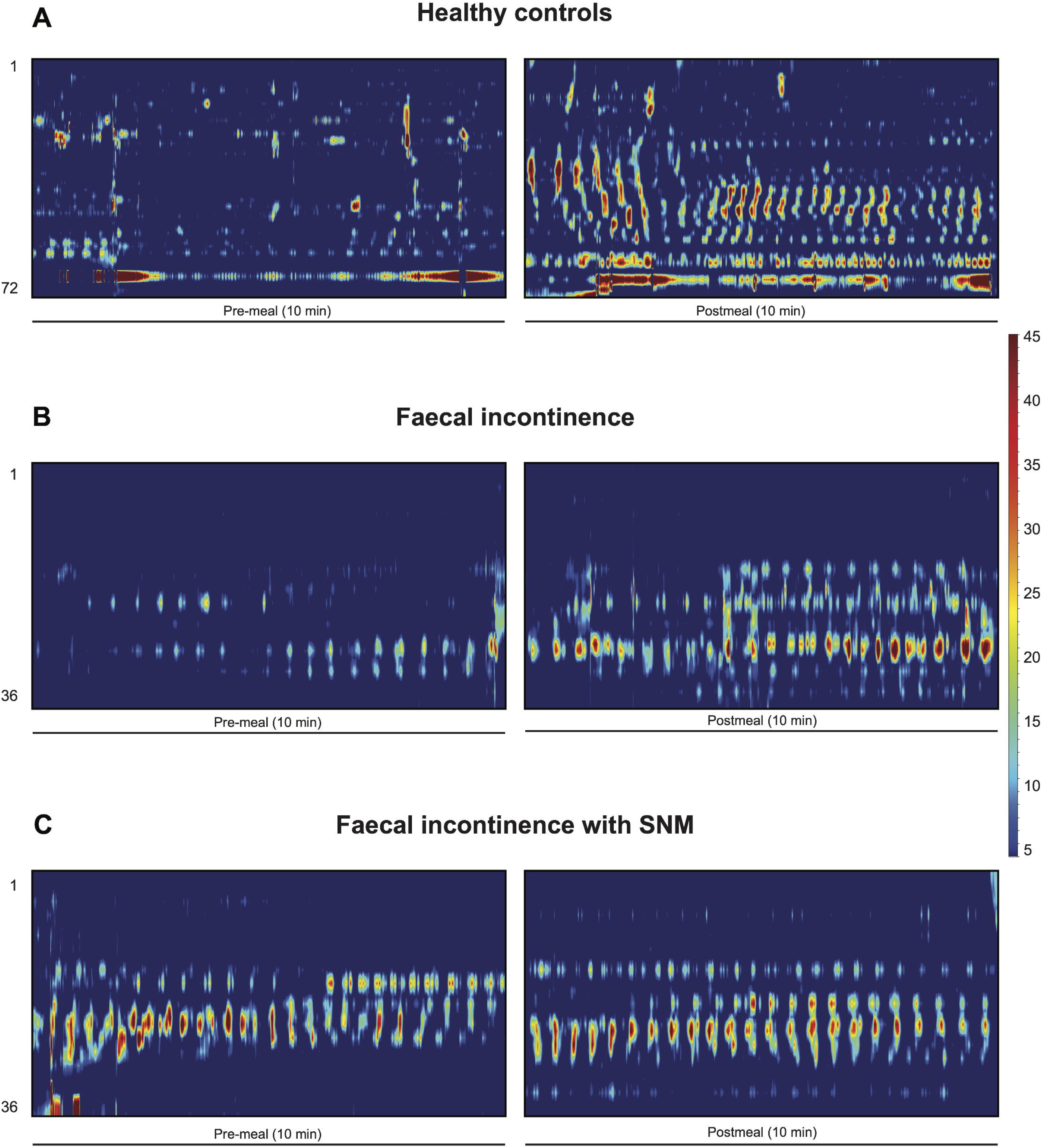
Effect of stimulation on the cyclic motor pattern stratified by meal and stimulation status. Plot depicts median (IQR); Paired nonparametric Wilcoxon test between pre-SNM and full-SNM comparisons: total p = 0.041, antegrade p = 0.264, retrograde p = 0.011.

**Figure 4:**
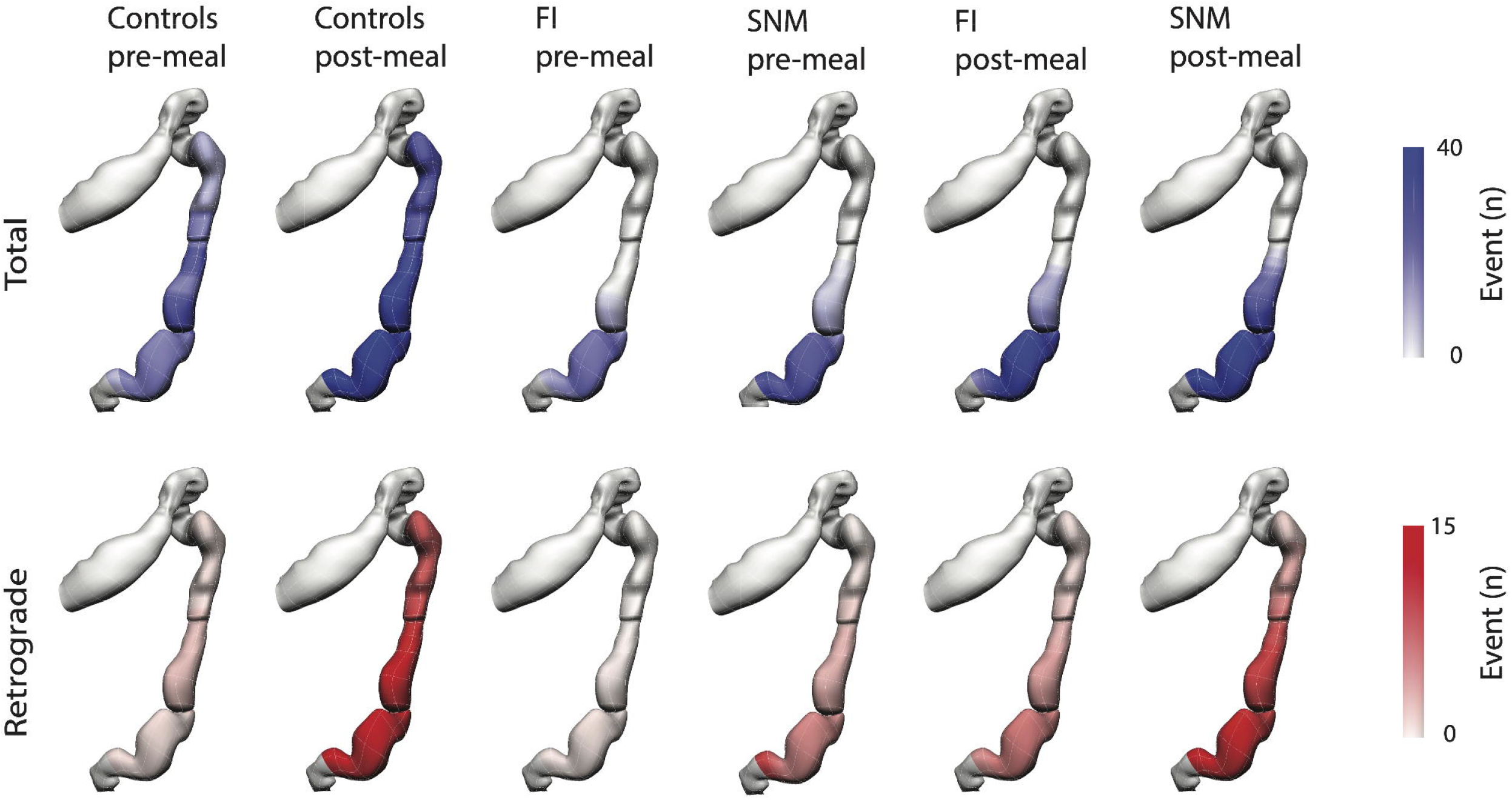
Anatomical registration of the event count distribution in a to a colonic geometry model, based on the estimated catheter insertion position. The colors represent the mean number of propagating events per hour. Propagating contractions were most active in the sigmoid colon. Total propagating contractions are depicted in blue and retrograde propagating contractions are depicted in red. FI, faecal incontinence; SNM, sacral neuromodulation.

### Effect of SNM on patients with faecal incontinence

SNM increased the number of propagating contractions compared to patients’ baselines. Among fasted patients with faecal incontinence, introduction of SNM increased the number of total (17 ± 6/h pre-meal vs 25 ± 5/h pre-meal with SNM, p = 0.043), retrograde (8 ± 3/h pre-meal vs 14 ± 3/h pre-meal with SNM, p = 0.028) propagating contractions (**Figure 1, 2 and 4**), but not antegrade propagating contractions (10 ± 3/h pre-meal vs 11 ± 4/h pre-meal with SNM, p = 0.527). Hence, SNM was shown to partially restore the meal response that was deficient in patients with FI compared to healthy controls (**Figure 2 and 4**).

SNM however did not fully restore the normal meal response. For example, in the fed state, there was no significant increase in total (22 ± 9/h post-meal without SNM vs 33 ± 8/h post-meal with SNM, p = 0.156), antegrade (5 ± 2/h post-meal without SNM vs 14 ± 3/h post-meal with SNM, p = 0.156) or retrograde (17 ± 9/h post-meal without SNM vs 19 ± 6/h post-meal with SNM, p = 0.313) propagating contractions with SNM (**Figure 2 and 4**). Therefore, SNM increased the baseline frequency of propagating contractions but not significantly more so in the fed state. Hence, if patients consumed a meal while stimulation was activated there was no significant further effect in the number of propagating contractions pre- and post-meal (p>0.05) (**Supplementary Appendix; Table S2**).

### Cyclic motor pattern in faecal incontinence and health

Similar to the primary analysis of all propagating contractions, patients with faecal incontinence showed an abnormal absence of the meal-response with respect to propagating contractions associated with the cyclic motor pattern. There was no increase in the number of total (11 ± 4/h pre-meal vs 12 ± 8/h post-meal, p = 0.423), antegrade (6 ± 2/h pre-meal vs 5 ± 4/h post-meal, p = 1.00), or retrograde (5 ± 2/h pre-meal vs 7 ± 4/h post-meal, p = 0.371) propagating contractions associated with the cyclic motor pattern (**Figure 1**). Healthy controls in contrast had significant increases in the number of total (16 ± 13/h pre-meal vs 87 ± 30/h post-meal, p = 0.010), and retrograde (9 ± 7/h pre-meal vs 68 ± 20/h post-meal, p = 0.003) propagating contractions (**Figure 1**), but not antegrade (7 ± 6/h pre-meal vs 19 ± 11/h post-meal, p = 0.361) propagating contractions.

### Effect of SNM on the cyclic motor pattern in patients with faecal incontinence

The effect of SNM on the number of propagating contractions associated with the cyclic motor pattern compared to baseline in patients with faecal incontinence did not reach significance. Among fasted patients with faecal incontinence, introduction of SNM resulted in a statistically insignificant increase in the number of total (11 ± 4/h pre-meal vs 17 ± 5/h pre-meal with SNM, p = 0.107), antegrade (6 ± 2/h pre-meal vs 8 ± 3/h pre-meal with SNM, p = 0.932), and retrograde (5 ± 2/h pre-meal vs 9 ± 4/h pre-meal with SNM, p = 0.080) propagating contractions associated with the cyclic motor pattern (**Figure 1**). However, when comparing the effect of SNM irrespective of meal status (i.e., full SNM compared to pre-SNM while amalgamating pre-meal and post-meal periods), significant increases in total and retrograde propagating contractions associated with the cyclic motor pattern were seen (p<0.05; **Figure 3**).

SNM however did not fully restore the normal meal response. For example, in the fed state, there was no significant increase in total (12 ± 8/h post-meal without SNM vs 20 ± 5/h post-meal with SNM, p = 0.098), antegrade (5 ± 4 post-meal without SNM vs 9 ± 3/h post-meal with SNM, p = 0.098) or retrograde (7 ± 4/h post-meal without SNM vs 11 ± 3/h post-meal with SNM, p = 0.098) propagating contractions with SNM. SNM therefore likely did not confer further enhancements to the rectosigmoid meal response when evaluating the cyclic motor pattern alone. However, only 6 patients had post-meal recordings without SNM.

### Clinical Outcomes

Two patients did not progress to a permanent SNM implant, one due to new onset of severe constipation after stage 1 SNM, and another due to <50% improvement in incontinence symptoms. The former patient with constipation had increased frequency of propagating activity at baseline and the latter had infrequent propagating contractions at baseline which slightly increased with SNM (**Supplementary Appendix; Figure S1**). Another patient required a change in the SNM programme prior to permanent implant placement. Median follow-up among 13 patients after second stage SNM was 47.0 (range 0.2-62.0) months. Eleven patients reported satisfaction with their bowel function whereas two patients reported deteriorating function, awaiting stimulator reprogramming.

## Discussion

Sacral neuromodulation is an effective treatment for faecal incontinence; however the mechanism of action has remained uncertain, limiting therapeutic progress. This study has shown that patients with faecal incontinence have an impaired rectosigmoid brake. Patients had fewer overall propagating contractions, and particularly retrograde propagating contractions, in response to a meal-stimulus in comparison to healthy controls. Secondly, this study has shown that SNM significantly increases the total number of propagating contractions in the rectosigmoid region, particularly retrograde propagating contractions, demonstrating a likely mechanism through which SNM exerts its therapeutic benefit.

There is consistent and advancing evidence of the importance of the rectosigmoid brake in the maintenance of normal continence ^12,19,20,29^. A ‘functional sphincter’ has long been recognised in this region, first attributed to O’Beirne ^30^. Chen et al. further characterized this ‘sphincter’ as an intermittent pressure band lying 10-17 cm above the anal verge, which relaxes and contracts in concert with the anal sphincters in response to pressure sequences ^31^. Dinning et al. applied HRCM to characterize a substantial postprandial increase in the retrograde cyclic motor pattern in the distal colon as a feature of a healthy meal response ^31^, and Lin et al. subsequently localised this activity to be maximal in the same rectosigmoid region as the ‘functional sphincter’ ^32^. Using another modality, high-resolution impedance manometry has shown gas insufflation of the sigmoid colon initiates retrograde cyclic motor patterns to limit gas transit to the rectum ^32^. These studies extended the earlier work of Rao and Welcher, who proposed periodic rectal motor activity served as an ‘intrinsic braking mechanism that prevents the untimely flow of contents’ ^33^. In addition, surgical resection of this region has recently been shown to contribute to a pathological absence of the meal response and symptoms of bowel dysfunction in patients with low anterior resection syndrome (LARS) ^34^. Based on these several background studies, we hypothesised that attenuation of rectosigmoid brake activity could also be an important pathophysiological mechanism of faecal incontinence. This hypothesis was confirmed in a cohort of severe medically-refractory incontinence patients in this study, as demonstrated by significantly reduced retrograde propagating activity in both fasting and fed states compared to controls.

Rectosigmoid motor activity has been shown to require neural innervation, as evidenced by its absence in spinal cord injury ^35,36^, systemic sclerosis ^37^, and diabetes mellitus ^38^, defining a plausible pathway through which SNM may act. Our data demonstrate that SNM effectively upregulates this pathway to effect enhanced retrograde distal colonic motility in patients with faecal incontinence, thereby confirming and extending previous work by Patton et al who first applied HRCM to demonstrate this effect ^19^. Further corroborating evidence for this effect has been provided by Michelson et al. using colorectal scintigraphy, who demonstrated that SNM decreased antegrade transit and increased retrograde transit in the descending colon, thereby prolonging colonic transit time and increasing colonic storage capacity ^42^. Together, these data now present a convincing body of evidence that modulation of colonic motility is a fundamental mechanism of action of SNM. Indeed, in light of the potent efficacy of SNM in many cases of anal sphincter incompetence ^11,40–42^, it can be posited that modulation of colonic motility, likely through the parasympathetic pelvic splanchnic nerves, may be a primary mechanism of action of this therapy, working in conjunction with other coregulatory phenomena such as potentially cortical activation ^43^.

The effect of a meal-response in the context of SNM has not previously been explored with HRCM. In this study, we found that a meal did not further significantly increase the number of propagating contractions beyond its baseline upregulating effect in patients with faecal incontinence (**Figure 2**), when compared with controls. This is similar to the findings of Roger et al. ^44^, who also found no difference in the frequency of manometric waves after a meal in patients with urge faecal incontinence. Notably, the study by Roger et al. employed low resolution pull-through colonic manometry, a technique that may miss a significant proportion of propagating sequences ^45^.

An expert consensus by Tack et al. recently proposed five criteria to qualify a putative pathophysiological mechanism in functional gastrointestinal disorders ^46^. Our findings of defective rectosigmoid brake activity in faecal incontinence can be usefully evaluated within this framework. Specifically, this study adds evidence to ‘Criterion 1’, which states that the pathophysiological disturbance is present in at least a subset of patients with the symptom, and the prevalence is higher than in appropriate controls. We also provide evidence for ‘Criterion 5’, which states that treatment, in this case through SNM, aimed at correcting the underlying disorder improves the symptom. This is evidenced by a partial restoration of the rectosigmoid brake with SNM, particularly in patients that benefited from SNM therapy. We did not investigate ‘Criterion 2’; whereby there should be a close temporal association between the pathophysiological disturbance and symptom occurrence, given that faecal incontinence is continuous. ‘Criterion 3’ states that there should be a significant correlation between the presence/severity of the symptom and the presence/severity of the dysfunction, and this criterion could be the focus of future work in a larger cohort of patients with broader range of symptom severities.

SNM has revolutionised the management of faecal incontinence. Over time, the threshold to offer SNM, which was once a last-line treatment option for medically-refractory patients, has been reduced ^8,47^. However, despite its success, the lack of actionable biomarkers for the efficacy of SNM has limited progress in advancing the therapy, for example in the 10-30% ^48^ of non-responders, and objective evaluation of stimulation protocols that could reduce energy consumption and prolong implant lifespan when optimised ^49^. The lack of a biomarker has also inhibited the development of less-invasive approaches, which would be applicable to a larger range of patients, such as tibial nerve stimulation ^50^. While this study shows that the rectosigmoid brake may offer a key biomarker, its assessment with HRCM is notably invasive, expensive, and analytically complex, limiting its broader utility. However, novel, non-invasive approaches to measure distal colonic motility are currently emerging, notably high-resolution electrocolonography ^51^.

As rectosigmoid brake is hypothesised to limit rectal filling, it is plausible that a hyperactive rectosigmoid brake may result in constipation in a subset of patients ^27^. While, the role of cyclic motor patterns in constipation is incompletely understood ^51–53^, some studies have demonstrated that increased retrograde rectosigmoid motility and pressure may impair bowel motions ^54^. Hyperactive retrograde rectosigmoid motility has also recently been shown to delay gut recovery after surgery ^54^, with bowel function not appearing to return until after rectosigmoid activity normalises ^55^. Interestingly, the patient in our study who failed to progress to permanent SNM due to onset of marked constipation after stage one had the highest frequency of retrograde cyclic motor patterns at baseline which increased further after SNM (**Figure S1**). In essence, SNM may therefore have resulted in overtreatment in a patient who did not suffer from rectosigmoid brake hypoactivity. A highly active rectosigmoid brake could therefore be one approach to help predict patients unlikely to respond to SNM in order to aid patient selection. However, further data is now needed to validate this hypothesis-generating observation from a single patient.

The present study has some limitations. Patients and controls received different bowel preparation, and patients also retained the manometric catheter in-situ for longer periods, which could hypothetically lead to a confounding effect of rectal filling ^56,57^. However, while bowel preparation alters the detection of HAPs and pre-defecatory motility patterns, it is not considered to alter the overall frequency of propagating contractions and the interpretation of meal responses as was the primary focus in this study ^58^. There were a relatively small number of patients in this study, reflecting the invasiveness of the technique and inconvenience for participating patients. Nevertheless, the data were sufficient to demonstrate statistically-robust effects for the primary hypothesis. The pathophysiology of incontinence is multifactorial, and this study included and analysed patients with both passive and urge incontinence together. Patients with both subtypes have shown improvements with SNM, however, there are likely underlying differences in pathophysiology, and potentially mechanism of response to SNM. However, it can also be argued that the consistency of findings in a diverse cohort strengthens this study’s conclusions.

In conclusion, patients with faecal incontinence are shown to have an impaired rectosigmoid brake, and attenuated postprandial increase in retrograde propagating contractions. SNM upregulates the rectosigmoid brake as evidenced by increased retrograde motility, likely aiding the maintenance of bowel continence. SNM did not, however, augment the meal response. The rectosigmoid brake is likely an important contributor to faecal continence and may represent a key biomarker for the effect of SNM.

## Data Availability

Data availability statement: The datasets generated during and/or analysed during the current study are not publicly available. Data sharing requests will be considered at the corresponding authors discretion.

## Acknowledgements

The authors would like to thank the patients and volunteers who contributed to this study.

## Supplementary Appendix

## Methods

Patients were positioned in a prone position with a pillow under the lower abdomen to flatten the sacrum. A grounding electrode was attached to either foot. After sterile preparation and draping, the S3 foramen location was identified using anatomical landmarks ^58^. Local anaesthetic (1% lidocaine) was infiltrated first if the procedure was carried out under sedation. A foramen needle was introduced at an approximately 60-degree angle into the S3 foramen. Typically, the left S3 foramen was accessed first. Fluoroscopy was used to confirm the position of the needle, both in the anterior-posterior and lateral orientation. The needle was then attached to an external test stimulator using a J-hook. A satisfactory position was indicated by the direct observation of the bellows response while minimising flexion of the great toe. If the response was unsatisfactory, the needle was repositioned on the ipsilateral side or contralateral side. Once a satisfactory position was confirmed, the foramen needle stylet was exchanged for guidewire followed by a rigid dilator within an introducer sheath through which the quadripolar tined lead (Model 3889, Medtronic, Minneapolis, MN, USA) was introduced. The position was confirmed using fluoroscopy and further test stimulation, looking for the bellows response. The lead was then tunneled subcutaneously to exit on the contralateral buttock, followed by standard skin closure. A sterile, waterproof dressing was applied. The electrode was then connected to a temporary external stimulator (Medtronic Verify Evaluation System, Model 3531) controlled by a patient (model 3537). The test lead was left disconnected after the procedure.

## Results

**Figure S1:**
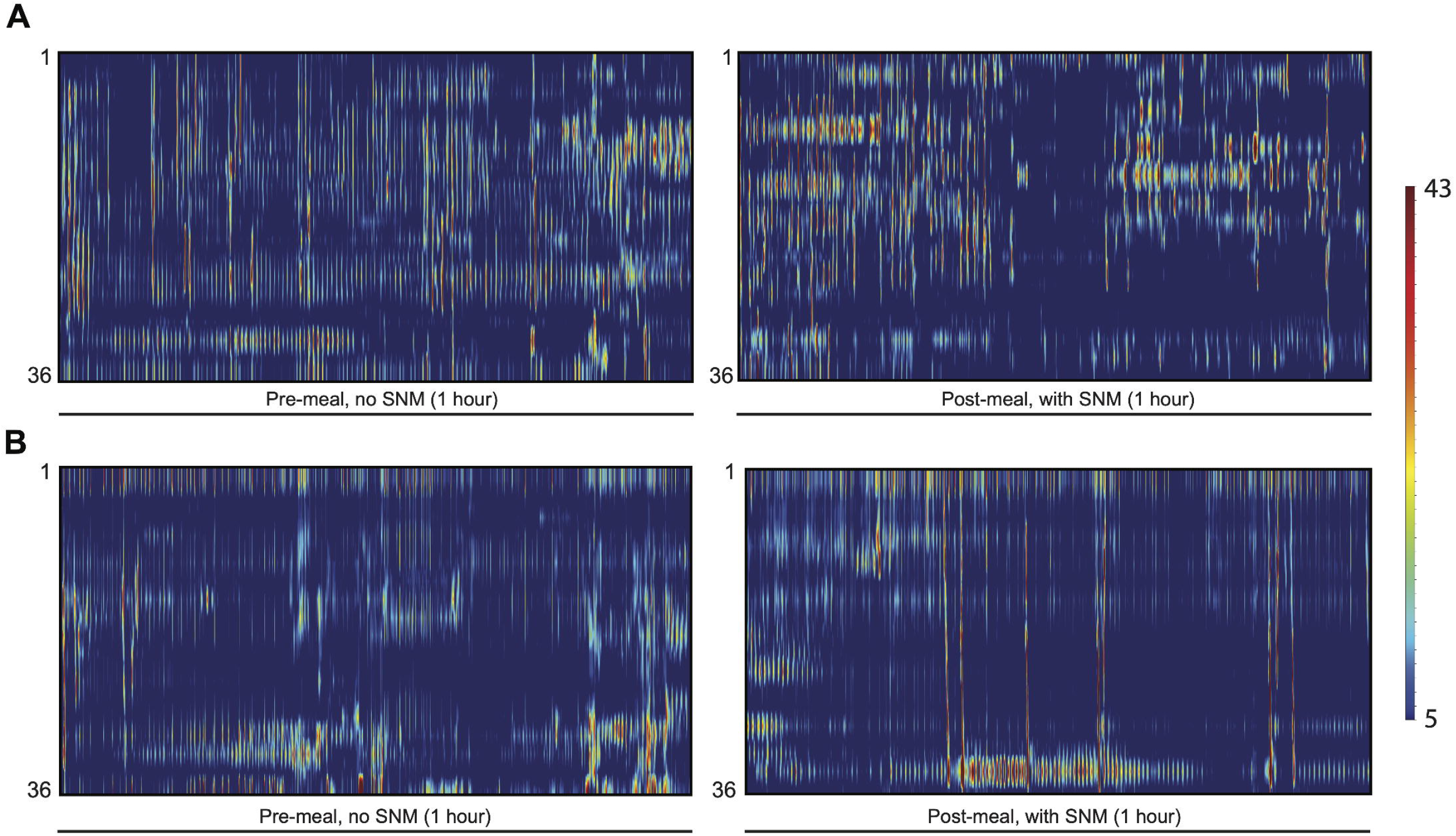
Manometry traces of patients that did not progress to stage 2 sacral neuromodulation. A: Manometric colour plots of the patient with faecal incontinence that had their stage 1 SNM removed secondary to constipation. Demonstrates increased levels of baseline (pre-meal, before SNM) propagating activity. B: Manometric colour plots of the patient with faecal incontinence that did not progress to stage 2 SNM due to suboptimal response during trial period.

**Table S1:**
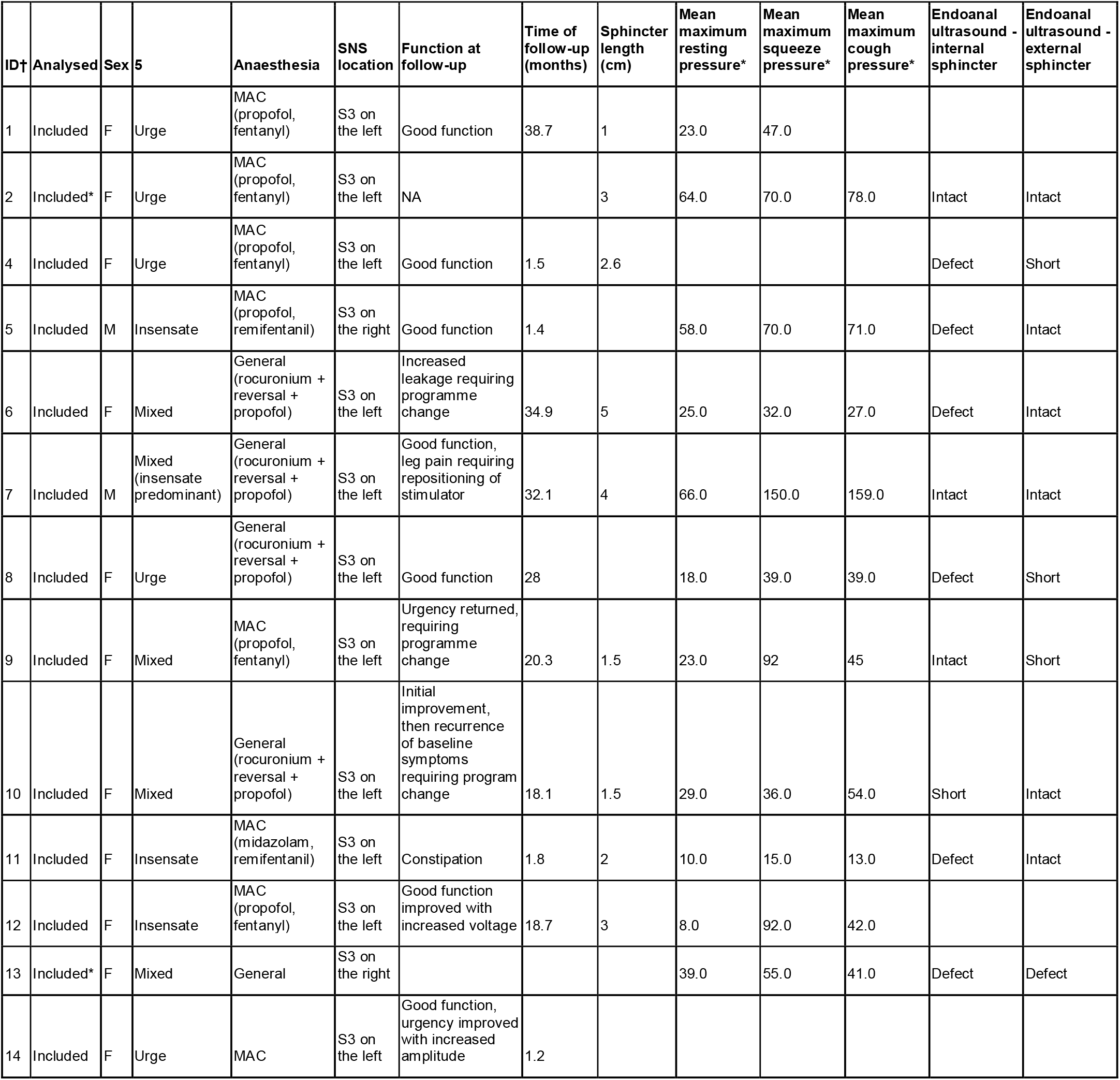
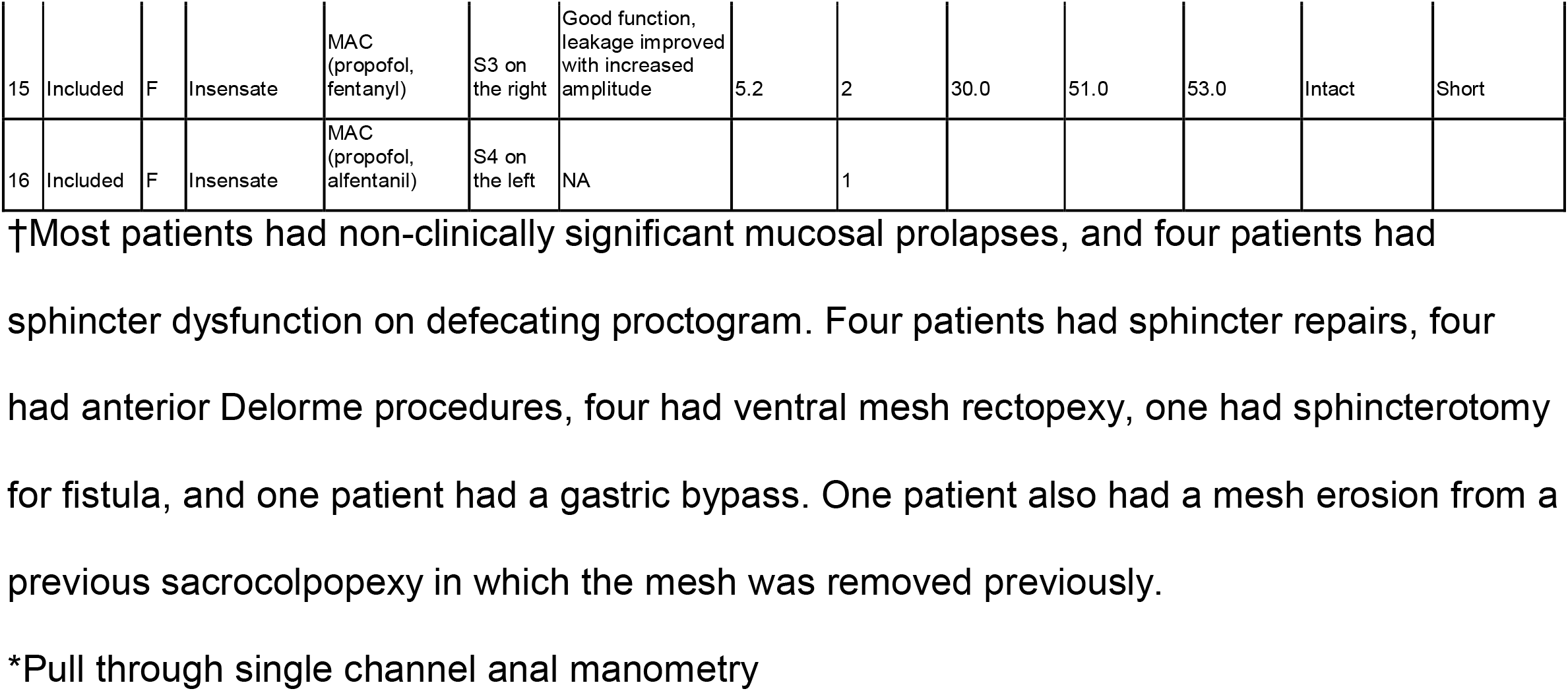
Clinical details of faecal incontinence cohort that had manometry analysed

| ID† | Analysed | Sex | 5 | Anaesthesia | SNS location | Function at follow-up | Time of follow-up (months) | Sphincter length (cm) | Mean maximum resting pressure* | Mean maximum squeeze pressure* | Mean maximum cough pressure* | Endoanal ultrasound - internal sphincter | Endoanal ultrasound - external sphincter |
| --- | --- | --- | --- | --- | --- | --- | --- | --- | --- | --- | --- | --- | --- |
| 1 | Included | F | Urge | MAC (propofol, fentanyl) | S3 on the left | Good function | 38.7 | 1 | 23.0 | 47.0 |  |  |  |
| 2 | Included* | F | Urge | MAC (propofol, fentanyl) | S3 on the left | NA |  | 3 | 64.0 | 70.0 | 78.0 | Intact | Intact |
| 4 | Included | F | Urge | MAC (propofol, fentanyl) | S3 on the left | Good function | 1.5 | 2.6 |  |  |  | Defect | Short |
| 5 | Included | M | Insensate | MAC (propofol, remifentanyl) | S3 on the right | Good function | 1.4 |  | 58.0 | 70.0 | 71.0 | Defect | Intact |
| 6 | Included | F | Mixed | General (rocuronium + reversal + propofol) | S3 on the left | Increased leakage requiring programme change | 34.9 | 5 | 25.0 | 32.0 | 27.0 | Defect | Intact |
| 7 | Included | M | Mixed (insensate predominant) | General (rocuronium + reversal + propofol) | S3 on the left | Good function, leg pain requiring repositioning of stimulator | 32.1 | 4 | 66.0 | 150.0 | 159.0 | Intact | Intact |
| 8 | Included | F | Urge | General (rocuronium + reversal + propofol) | S3 on the left | Good function | 28 |  | 18.0 | 39.0 | 39.0 | Defect | Short |
| 9 | Included | F | Mixed | MAC (propofol, fentanyl) | S3 on the left | Urgency returned, requiring programme change | 20.3 | 1.5 | 23.0 | 92 | 45 | Intact | Short |
| 10 | Included | F | Mixed | General (rocuronium + reversal + propofol) | S3 on the left | Initial improvement, then recurrence of baseline symptoms requiring program change | 18.1 | 1.5 | 29.0 | 36.0 | 54.0 | Short | Intact |
| 11 | Included | F | Insensate | MAC (midazolam, remifentanyl) | S3 on the left | Constipation | 1.8 | 2 | 10.0 | 15.0 | 13.0 | Defect | Intact |
| 12 | Included | F | Insensate | MAC (propofol, fentanyl) | S3 on the left | Good function improved with increased voltage | 18.7 | 3 | 8.0 | 92.0 | 42.0 |  |  |
| 13 | Included* | F | Mixed | General | S3 on the right |  |  |  | 39.0 | 55.0 | 41.0 | Defect | Defect |
| 14 | Included | F | Urge | MAC | S3 on the left | Good function, urgency improved with increased amplitude | 1.2 |  |  |  |  |  |  |
| 15 | Included | F | Insensate | MAC (propofol, fentanyl) | S3 on the right | Good function, leakage improved with increased amplitude | 5.2 | 2 | 30.0 | 51.0 | 53.0 | Intact | Short |
| 16 | Included | F | Insensate | MAC (propofol, alfentanil) | S4 on the left | NA |  | 1 |  |  |  |  |  |

†Most patients had non-clinically significant mucosal prolapses, and four patients had sphincter dysfunction on defecating proctogram. Four patients had sphincter repairs, four had anterior Delorme procedures, four had ventral mesh rectopexy, one had sphincterotomy for fistula, and one patient had a gastric bypass. One patient also had a mesh erosion from a previous sacrocolpopexy in which the mesh was removed previously.
\*Pull through single channel anal manometry

**Table S2:** Meal response in faecal incontinence with full strength SNM activated

| Metric (propagating contractions) | Pre-meal (1 h) | Post-meal (1 h) | p |
| --- | --- | --- | --- |
| Total | 25 ± 5 | 33 ± 8 | 0.272 |
| Antegrade | 11 ± 4 | 14 ± 3 | 0.414 |
| Retrograde | 14 ± 3 | 19 ± 6 | 0.552 |

